# Predictive targeting of mitochondrial metabolism in Acute Myeloid Leukemia patients with a lipoic acid analog

**DOI:** 10.1101/2021.06.03.21257935

**Authors:** Michael A. Reid, Shree Bose, Kristin M Pladna, Rebecca Anderson, Peter G. Mikhael, Zhengtao Xiao, Ziwei Dai, Shiyu Liu, Juan Liu, Timothy S. Pardee, Jason W. Locasale

**Author notes:** Correspondence to Jason W. Locasale – or Timothy S. Pardee. co-first authors (Michael A. Reid, Shree Bose).

## Abstract

Targeting metabolism has long been a theory for cancer therapy, but clinical development has been limited by toxicities, compound availability, overall efficacy, and patient specificity^1^. CPI-613, a lipoic acid analogue that interferes with enzymes involved in mitochondrial metabolism, has demonstrated clinical activity in lethal malignancies including relapsed or therapy refractory Acute Myeloid Leukemias (AMLs)^2,3^ and Phase III trials are ongoing^1^. Using metabolomics, we investigated blood and bone marrow samples from a cohort of 29 relapsed or refractory AML patients involved in Phase I and II studies undergoing CPI-613 treatment (NCT01768897, NCT02484391) including 13 that achieved a complete response. We show that CPI-613 treatment in patients induced defined alterations related to the tricarboxylic acid (TCA) cycle and associated redox, anabolic and catabolic metabolism. These findings are consistent with targeting of several ketoacid dehydrogenase (KADH) enzymes that use lipoic acid as a cofactor and are related to mitochondrial metabolism. The alterations were observed systemically but were more pronounced within the leukemic bone marrow microenvironment consistent with its mechanistic target. Machine learning revealed that metabolic status and changes associated with mitochondrial metabolism were predictive of treatment response, indicating that mechanism-based metabolite biomarkers to a targeted metabolic cancer therapy may be feasible. Finally, we confirm using isotope tracing and flux analysis that these effects are due to disruptions to substrate utilization into the mitochondria. Our findings provide evidence that a tolerated, anti-cancer therapeutic can act by targeting mitochondrial metabolism in humans.

## INTRODUCTION

Cancers adapt metabolism to meet the demands of energy, redox maintenance, biosynthesis, signaling, chromatin maintenance among other functions^4^. Many of these requirements are met in the mitochondria by the usage of nutrients that feed the tricarboxylic acid (TCA) cycle from glucose, lipids and amino acids that is together referred to as central carbon metabolism. Substrate utilization in the TCA cycle as with all metabolic processes is plastic and depends on genetic status, metabolic network status as determined by enzyme expression and activity, and particularly nutrient availability in the environment ^5,6^. These differential metabolic requirements of cancer associated mitochondria and central carbon metabolism have long been hypothesized as targetable vulnerabilities in cancer and pre-clinical studies show promise^1,7-9^. However, the TCA cycle is required for function in nearly every healthy cell and conserved to the most primitive of organisms. Thus, successfully targeting metabolism in cancer by defining its window that satisfies both efficacy and toxicity is complex.

Despite the numerous potential targets accumulated from thousands of papers and substantial commercial efforts, the establishment of clinical-stage therapies targeting cancer cell metabolism in humans has been limited. CPI-613 or Devimistat^10^, a lipoic acid analogue has potentially intriguing properties in relation to cancer metabolism due to the family of enzymes involved in mitochondrial metabolism that use lipoic acid as a cofactor^11^. In early phase human trials, CPI-613 has demonstrated substantial activity and tolerability with relapsed or refractory acute myeloid leukemia^2,12^ and pancreatic cancer^3^, two of the most aggressively lethal malignancies. Large-scale randomized, controlled phase 3 trials are ongoing^13,14^. However, whether the positive clinical results thus far are due to target engagement and subsequent alterations to mitochondrial metabolism in tumors is unknown. Thus despite this interesting clinical activity, it remains unclear whether cancer metabolism may be targeted in this fashion in humans.

## RESULTS

### The lipoic acid analogue CPI-613 alters blood plasma metabolite profiles in AML patients

Ketoacid dehydrogenase (KADH) enzyme complexes are trimers that consist of E1, E2, and E3 subunits that involve a set of enzymes related to metabolic reactions in the TCA cycle^11,15^. While the E1 subunit provides substrate-specificity, the E2 and E3 subunits are conserved^11^ and the E2 subunit requires lipoic acid to function via a covalent modification (Fig. 1a). Since CPI-613 is a lipoic acid analogue and thus influences the assembly and activity of KADH enzyme complexes (Fig. 1b) that are involved in central carbon metabolism^10,12,2,3^, we investigated the effects on metabolism of CPI-613 treatment on humans. We first performed metabolite profiling of blood plasma from a cohort of 29 AML patients undergoing a phase I or II study of CPI-613 in combination with high dose cytarabine and mitoxantrone collected at baseline and 1-6 days post CPI-613-dosing using liquid chromatography coupled to high-resolution mass spectrometry (LC-HRMS) (Fig. 1c). In samples of patient blood plasma, we detected CPI-613 at concentrations after treatment consistent with previous pharmacokinetic reports which was importantly undetectable in the untreated patients^12,2^ (Fig. 1d, Supplementary Fig. 1). The concentrations did not differ between responders and non-responders indicating that response was not due to interindividual pharmacology or effective dosage (Fig. 1e). Moreover, CPI-613 treatment overall produced a defined metabolic signature (Fig. 1f) following CPI-613 treatment in patient blood plasms that corresponded to metabolic pathways putatively affected by CPI-613 based on where the enzymes that utilize lipoic acid sit in the metabolic network (Fig. 1g, h). Together, these results indicate CPI-613 treatment results in alterations to metabolism in human AML patients with features related to metabolism involved in lipoic acid utilizing enzymes.

**Figure 1.**
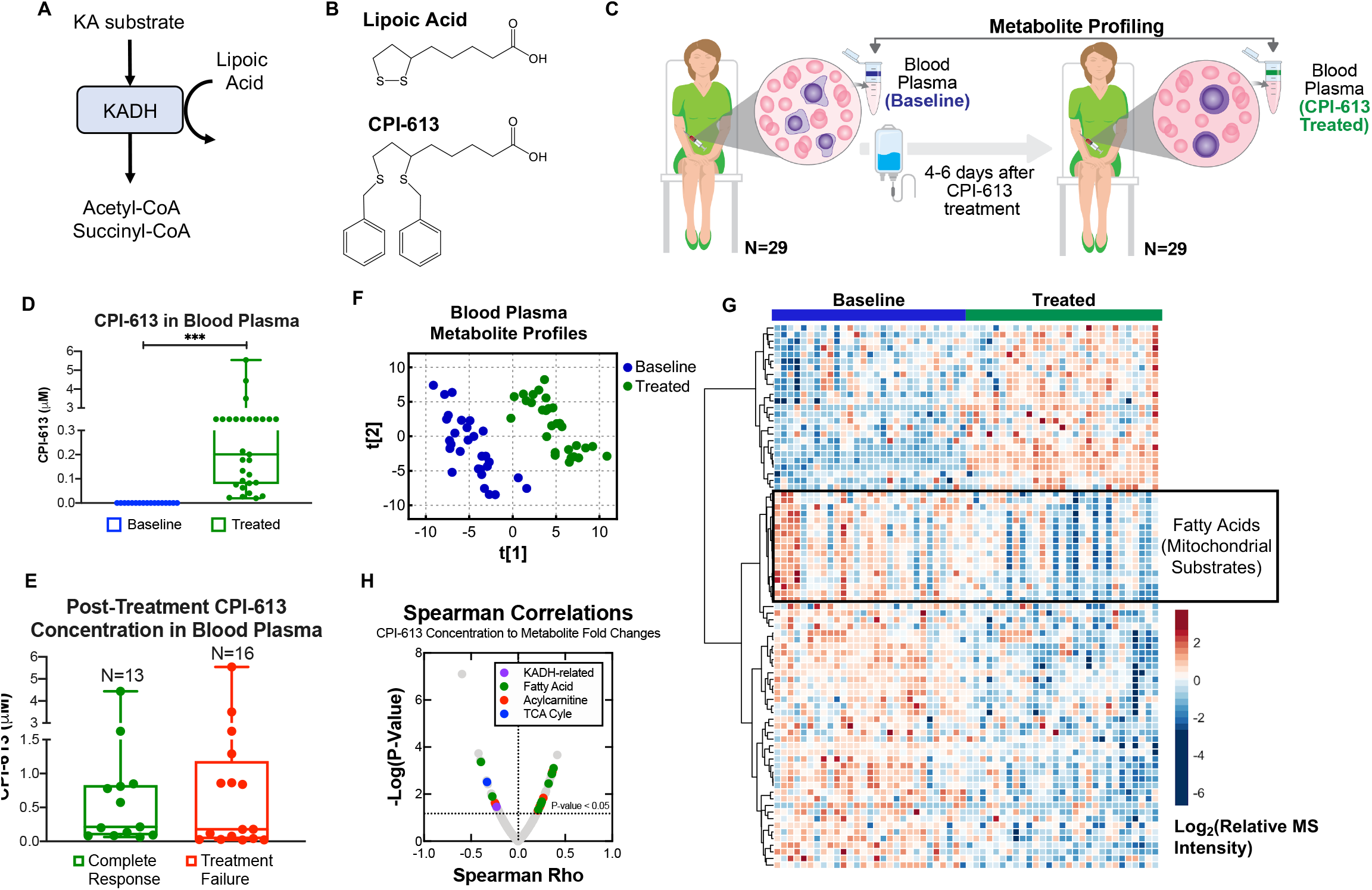
The lipoic acid analogue CPI-613 alters blood plasma metabolite profiles in AML patients. (a) Schematic of ketoacid dehydrogenase (KADH) enzymatic activity. Ketoacid (KA) substrates are converted to acetyl-coA or succinyl-coA through a sequence of events that require lipoic acid. (b) Chemical structures of lipoic acid and CPI-613. (c) Schematic of the acute myeloid leukemia patient blood plasma sample collection procedure at baseline and 4-6 days after CPI-613 treatment. Metabolite profiling was performed on blood plasma from 29 AML patients at baseline and after CPI-613. (d) Absolute quantitation of CPI-613 (μM) in AML patient blood plasma following CPI-613 treatment. The upper and lower whiskers of the box plot are the maximum and minimum values, respectively, and the bar is the median value. Data are from 29 AML patients, and individual values are plotted. *P* < 0.005 [***], paired Student’s *t*-test. (e) Absolute quantitation of CPI-613 (μM) in AML patient blood plasma following CPI-613 treatment stratified by patient outcome. The upper and lower whiskers of the box plot are the maximum and minimum values, respectively, and the bar is the median value. Data are from 29 AML patients, and individual values are plotted. (f) Partial least squares-discriminant analysis (PLS-DA) was performed to examine the variance between blood plasma metabolite profiles of individual patients before and after treatment. Blue dots represent individual patients at baseline and green dots represent individual patients after CPI-613 treatment. (g) Heat map of metabolite profiles from individual AML patients’ blood plasma before and after CPI-613 treatment. (h) Spearman correlation of blood plasma CPI-613 concentration to metabolite fold changes.

### Blood plasma profiling reveals that CPI-613 affects KADH activity and mitochondrial metabolism in patients

To investigate target engagement, we examined whether CPI-613 treatment affected KADH enzyme activity in AML patients. KADH substrate source levels are known to be indicative of KADH enzyme functionality. For example, humans with inborn errors in the gene encoding Lipoyltransferase-1, a key component in lipoic acid metabolism, display reduction of KADH enzyme function and accumulation of KADH substrate sources in blood plasma^16^. Thus, we assayed all KADH substrate sources in the AML cohort blood plasma at baseline and after CPI-613 treatment (Fig. 2a). Consistent with the reduction of KADH enzyme activity by CPI-613, we observed increases in KADH substrate source levels in treated blood plasma (Fig. 2b). At the individual patient level, we found accumulation of certain KADH substrates such as the branched chain amino acid valine in up to 21 of the 29 patients after CPI-613 treatment (Fig. 2c). Furthermore, each patient showed reduced activity in at least one of the KADH-family enzymes after CPI-613 treatment (Supplementary Fig. 2a). Since KADH-family enzymes promote mitochondrial metabolism^11^, we sought to determine whether CPI-613 had any effect on the TCA cycle and related metabolism in AML patients. The levels of blood plasma acylcarnitines, which are derived from fatty acids, are known to be direct read-outs of cellular mitochondrial status^17,18^. Of significance, we found AML patients treated with CPI-613 had alterations in the levels of acylcarnitines (Fig. 2d) and fatty acids (Supplementary Fig. 2b). In fact, many metabolites differing in magnitude after treatment in patient blood plasma compared to baseline were related to ketoacid dehydrogenase and mitochondrial metabolism in both responders (CR; Complete Remission) and non-responders (TF; Treatment Failure) (Fig. 2e,f). In all, these results demonstrate that CPI-613 treatment leads to systemic reduction of KADH enzyme activity and altered mitochondrial metabolism in AML patients confirming its target in humans.

**Figure 2.**
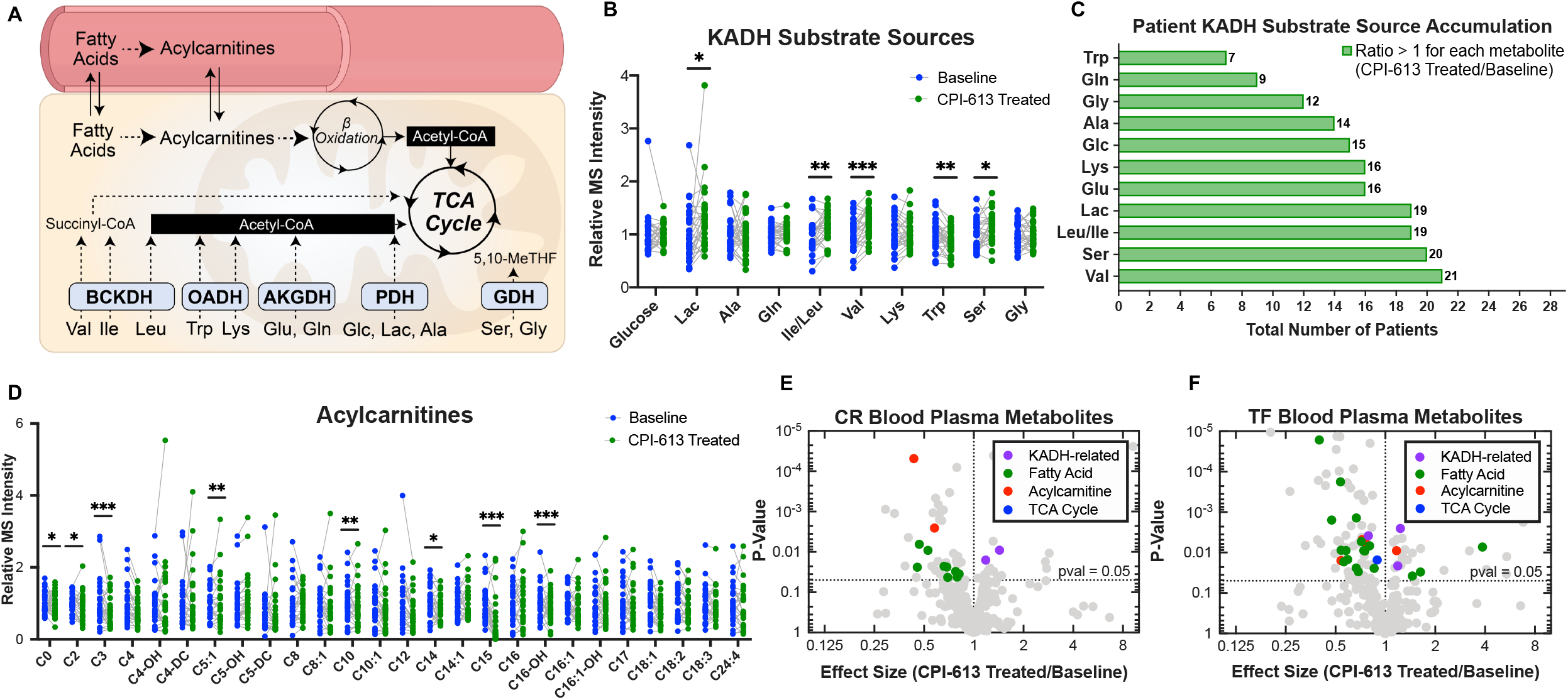
CPI-613 affects KADH enzymatic activity and mitochondrial metabolism in AML patients. (a) Schematic of ketoacid dehydrogenase (KADH) substrate source inputs and their biochemical output via KADH enzymatic activity. PDH, pyruvate dehydrogenase; OADH, 2-oxoacid dehydrogenase; AKGDH, alpha-ketoglutarate dehydrogenase; BCKDH, branched chain ketoacid dehydrogenase; GDH, glycine dehydrogenase; Glc, glucose; Lac, lactate; Ala, alanine; Trp, tryptophan; Lys, lysine; Glu, glutamate; Gln, glutamine; Val, valine; Ile, isoleucine; Leu, leucine; Ser, serine; Gly, glycine; TCA cycle, tri-carboxylic acid cycle. (b) Relative mass spectrometry (MS) peak intensities of KADH substrate sources in AML patient blood plasma at baseline and after CPI-613 treatment. The upper and lower whiskers of the box plots are the maximum and minimum values, respectively, and the bars are the median value. Data are from 29 AML patients, and individual values are plotted. *P* < 0.05 [*], *P* < 0.01 [**], *P* < 0.005 [***], paired Student’s *t*-test. (c) Total number of patients [out of 29] with indicated KADH substrate accumulation as determined by a ratio of >1 in the MS peak intensity measured in the CPI-613 treated blood plasma versus baseline in individual patients. (d) Relative mass spectrometry (MS) peak intensities of acylcarnitines in AML patient blood plasma at baseline and after CPI-613 treatment. The upper and lower whiskers of the box plots are the maximum and minimum values, respectively, and the bars are the median value. Data are from 29 AML patients, and individual values are plotted. *P* < 0.05 [*], *P* < 0.01 [**], *P* < 0.005 [***], paired Student’s *t*-test. (e) Volcano plots of blood plasma metabolite profiles in patients with complete remission (CR) responses to CPI-613. Data are mean of patient integrated peak intensities (Baseline n=13 patients; CPI-613 treated n=13 patients). Purple dots, ketoacid dehydrogenase (KADH)-related metabolites; Green dots, fatty acids; Red dots, acylcarnitines; Blue dots, TCA cycle metabolites. The dots represent metabolites that significantly changed between Baseline and CPI-613 treated patients (p<0.05). (f) Volcano plots of blood plasma metabolite profiles in patients with treatment failure (TF) responses to CPI-613. Data are mean of patient integrated intensities (Baseline n=16 patients; CPI-613 treated n=16 patients). Purple dots, ketoacid dehydrogenase (KADH)-related metabolites; Green dots, fatty acids; Red dots, acylcarnitines; Blue dots, TCA cycle metabolites. The dots represent metabolites that significantly changed between Baseline and CPI-613 treated patients (p<0.05).

### CPI-613 treatment outcome corresponds with bone marrow metabolite profiles

We then asked whether AML patient metabolite profiles corresponded to CPI-613 treatment outcome. While the comparison of blood plasma metabolite profiles revealed responders (CR) and non-responders (TF) had similar global alterations in KADH and mitochondrial-related metabolites after CPI-613 treatment, these observations confirmed systemic target engagement. As the niche of hematopoietic stem cells, the bone marrow is the major source of leukemic cells^19^. Thus, we reasoned that metabolite profiles of the bone marrow plasma that bathes the site of the tumor could provide deeper insight into the mechanism of CPI-613 action in AML patients. Accordingly, we performed LC-HRMS on patient baseline and nadir bone marrow plasma to examine this question (Fig. 3a). Indeed, distinct metabolite profiles were observed in the bone marrow of responders treated with the drug compared to baseline (Fig. 3b,c). Moreover, CPI-613 led to accumulation of KADH substrate sources and alterations in fatty acid and acylcarnitine levels in bone marrow plasma of complete responders (CR) (Fig. 3d, Supplementary Fig. 3), but not in the treatment failure (TF) cohort (Fig. 3e, Supplementary Fig. 4). We then directly examined whether CPI-613 affected mitochondrial metabolism in the tumor microenvironment by profiling the TCA cycle in the bone marrow plasma. Strikingly, we observed robust reduction of all TCA cycle intermediates in the CR patients (Fig. 3f), but not in TF patients (Fig. 3g). In all, while bone marrow plasma profiles of the CR cohort were consistent with overall effects on ketoacid dehydrogenase and mitochondrial metabolism in response to CPI-613 treatment (Fig. 3h), the TF cohort displayed almost no metabolic alterations (Fig. 3i). Collectively, these results provide evidence that the efficacy of CPI-613 occurs through engagement of KADH enzymes and disruption to mitochondrial metabolism, and that patient outcome is dependent on the capacity of the drug to alter mitochondrial metabolism at the tumor site.

**Figure 3.**
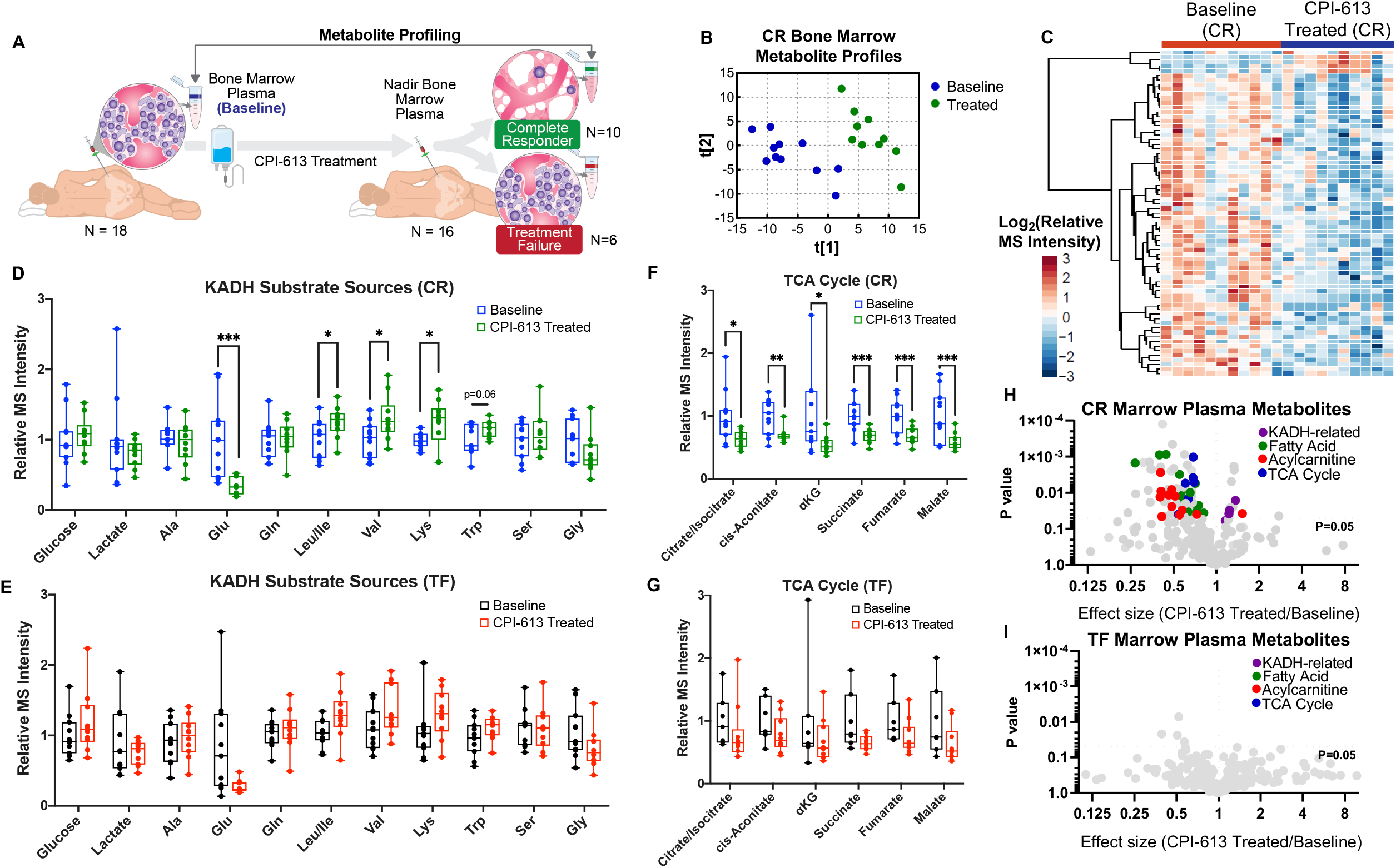
CPI-613 treatment outcome corresponds with bone marrow metabolite profiles. (a) Schematic of the acute myeloid leukemia patient bone marrow plasma sample collection procedure at baseline and after CPI-613 treatment (nadir bone marrow). Metabolite profiling was performed on bone marrow plasma from 18 AML patients at baseline and 16 AML patients after CPI-613. (b) Partial least squares-discriminant analysis (PLS-DA) was performed to examine the variance between bone marrow plasma metabolite profiles of individual complete remission (CR) patients before and after treatment. Blue dots represent individual patients at baseline and green dots represent individual patients after CPI-613 treatment. (c) Heat map of metabolite profiles from individual CR AML patients’ bone marrow plasma before and after CPI-613 treatment. (d) Relative mass spectrometry (MS) peak intensities of KADH substrate sources CR AML patient bone marrow plasma at baseline and after CPI-613 treatment. The upper and lower whiskers of the box plots are the maximum and minimum values, respectively, and the bars are the median value. Data are from CR AML patients (Baseline n=11 patients; CPI-613 treated n=10 patients), and individual values are plotted. *P* < 0.05 [*], *P* < 0.005 [***], unpaired Student’s *t*-test. Ala, alanine; Trp, tryptophan; Lys, lysine; Glu, glutamate; Gln, glutamine; Val, valine; Ile, isoleucine; Leu, leucine; Ser, serine; Gly, glycine. (e) Relative mass spectrometry (MS) peak intensities of ketoacid dehydrogenase (KADH)-related metabolites in treatment failure (TF) AML patient bone marrow plasma at baseline and after CPI-613 treatment. The upper and lower whiskers of the box plots are the maximum and minimum values, respectively, and the bars are the median value. Data are from AML patients (Baseline n=7 patients; CPI-613 treated n=6 patients). Ala, alanine; Trp, tryptophan; Lys, lysine; Glu, glutamate; Gln, glutamine; Val, valine; Ile, isoleucine; Leu, leucine; Ser, serine; Gly, glycine. (f) Relative mass spectrometry (MS) peak intensities of tri-carboxylic acid (TCA) cycle metabolites in CR AML patient bone marrow plasma at baseline and after CPI-613 treatment. The upper and lower whiskers of the box plots are the maximum and minimum values, respectively, and the bars are the median value. Data are from CR AML patients (Baseline n=11 patients; CPI-613 treated n=10 patients), and individual values are plotted. *P* < 0.05 [*], *P* < 0.01 [**], *P* < 0.005 [***], unpaired Student’s *t*-test. αKG, alpha-ketoglutarate. (g) Relative mass spectrometry (MS) peak intensities of tri-carboxylic acid (TCA) cycle metabolites in treatment failure (TF) AML patient bone marrow plasma at baseline and after CPI-613 treatment. The upper and lower whiskers of the box plots are the maximum and minimum values, respectively, and the bars are the median value. Data are from AML patients (Baseline n=7 patients; CPI-613 treated n=6 patients). αKG, alpha-ketoglutarate. (h) Volcano plots of bone marrow plasma metabolite profiles in patients with complete remission (CR) responses to CPI-613. Data are mean of patient integrated peak intensities (Baseline n=11 patients; CPI-613 treated n=10 patients). Purple dots, ketoacid dehydrogenase (KADH)-related metabolites; Green dots, fatty acids; Red dots, acylcarnitines; Blue dots, TCA cycle metabolites. The dots represent metabolites that significantly changed between Baseline and CPI-613 treated patients (p<0.05). (i) Volcano plots of bone marrow plasma metabolite profiles in patients with treatment failure (TF) responses to CPI-613. Data are mean of patient integrated peak intensities (Baseline n=7 patients; CPI-613 treated n=6 patients). Purple dots, ketoacid dehydrogenase (KADH)-related metabolites; Green dots, fatty acids; Red dots, acylcarnitines; Blue dots, TCA cycle metabolites. The dots represent metabolites that significantly changed between Baseline and CPI-613 treated patients (p<0.05).

### Metabolite concentrations in bone marrow plasma prior to CPI-613 administration can predict treatment outcome

The connection between bone marrow plasma metabolite profiles and AML patient response to CPI-613 was intriguing and warranted further investigation, particularly as to whether any predictive capacity of patient response could be gleaned from metabolic status. We thus investigated whether baseline metabolic phenotypes could stratify patient response and potentially predict treatment outcomes. To conduct this task, we considered a feature selection process involving metabolites with high mutual information, validated mass spectra, and known biological mechanism pursuant to mitochondrial metabolism (Fig 4a). We first performed two analyses involving computation of correlation metrics of baseline bone marrow plasma metabolite profiles. Interestingly, metabolites associated with KADH enzyme activity and mitochondrial metabolism (succinyl carnitine, dihydroorotate^20^ and aspartate^21^) were among the top metabolites linked to treatment outcome in both a Spearman correlation (Fig. 4b) and mutual information analysis (Fig. 4c). To explore the predictive capability of baseline bone marrow plasma metabolic profiles, a Lasso logistic regression model approach was used to determine the scaled Lasso coefficients for each of these 3 metabolites as well as other sets of metabolites fitting our criteria (Fig. 4d). Then, we generated ROC curves to determine the predictive capabilities of these metabolites. These calculations revealed that sets of three metabolites related to target engagement around mitochondrial metabolism were predictive of response (AUC = 0.71 to 0.93) (Fig. 4e). Interestingly, the combination of aspartate, dihydroorotate, and succinyl-carnitine yielded the greatest predictive capacity with an AUC of 0.93 (Fig. 4e,f). These results indicated inherent metabolic status related to mitochondrial metabolism within the tumor microenvironment predicts response to CPI-613.

**Figure 4.**
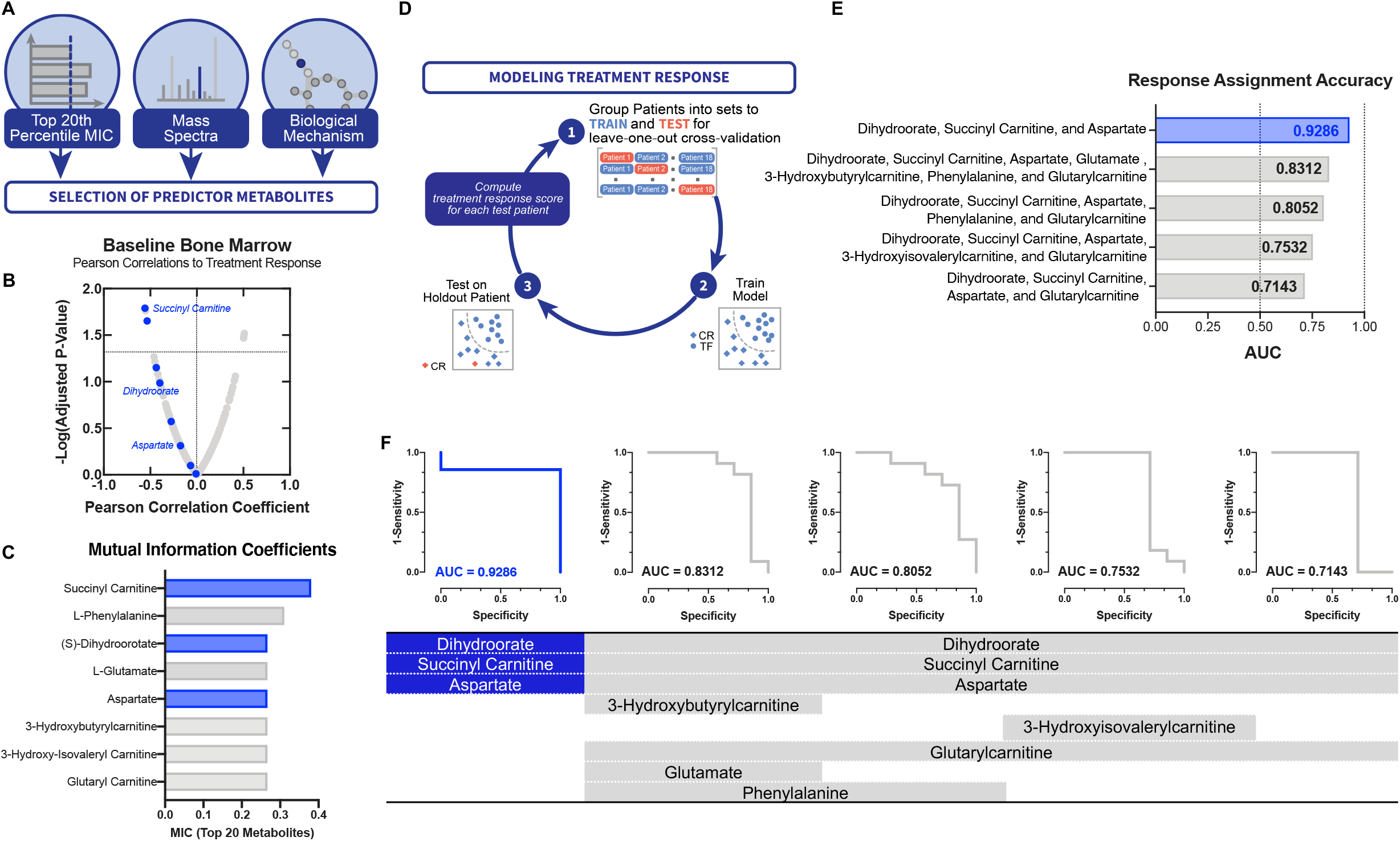
Metabolite concentrations in bone marrow plasma prior to CPI-613 administration can predict treatment outcome. (a) Schematic for metabolite feature selection. (b) Spearman correlation of baseline bone marrow plasma metabolic profiles of all patients to CPI-613 treatment response (CR vs. TF). (c) High ranking mutual information coefficients between metabolite level and CPI-613 treatment response. (d) Response assignment accuracy. (e) Accuracy of determining CPI-613 treatment outcome from bone marrow plasma metabolite levels of selected metabolites.

### CPI-613 reduces KADH activity and mitochondrial metabolic flux in human cells

Plasma metabolite profiles were indispensable for defining metabolic component and predictive capacity of treatment in human AML patients. Laboratory settings however provide avenues to model the human biology and test the theories obtained from their observations. We next sought to determine the effects of CPI-613 on cellular metabolism in a laboratory model. Of significance, cells treated with CPI-613 in culture had metabolite profiles indicative of KADH enzyme activity and mitochondrial metabolism inhibition (Fig. 5a). Indeed, metabolic pathway analysis revealed that the pathways most affected by CPI-613 treatment were related to amino acid and mitochondrial metabolism (Fig. 5b). Of particular interest, CPI-613 treated cells displayed metabolite profiles consistent with those observed in the blood and bone marrow plasma of AML patients treated with the drug including accumulation of KADH substrate sources (Fig. 5c), alterations in acylcarnitine levels (Supplementary Fig. 5a,b), and reduced levels of fatty acids (Supplementary Fig. 5c) and TCA cycle intermediates (Supplementary Fig. 5d). While total levels of metabolites yield insights into metabolic status, these profiles are not always sufficient for understanding metabolic activity which is characterized by flux through a given pathway^22,23^. Thus, we performed kinetic flux profiling of cells in the presence or absence of CPI-613. To do this, we cultured cells in the presence of DMSO or CPI-613 for 24h in ^12^C glucose-containing medium, then grew cells in [U-^13^C] glucose-containing medium (with the same treatments) and extracted metabolites at numerous time points. We then used LC-HRMS and examined enrichment of the [U-^13^C] glucose-derived m+2 isotopomers of the TCA cycle intermediates citrate/isocitrate, α-ketoglutarate, succinate, fumarate, and malate over time (Fig. 5d). We found that CPI-613 treatment resulted in significant reduction of metabolic flux through the TCA cycle (Fig. 5e-i). Indeed, quantitative metabolic flux analysis confirmed glucose flux through the KADH-family enzyme pyruvate dehydrogenase was significantly reduced (Fig. 5j,k). Taken together, these results confirm that CPI-613 targets KADH enzyme activity and reduces mitochondrial metabolism in human cancer cells.

**Figure 5.**
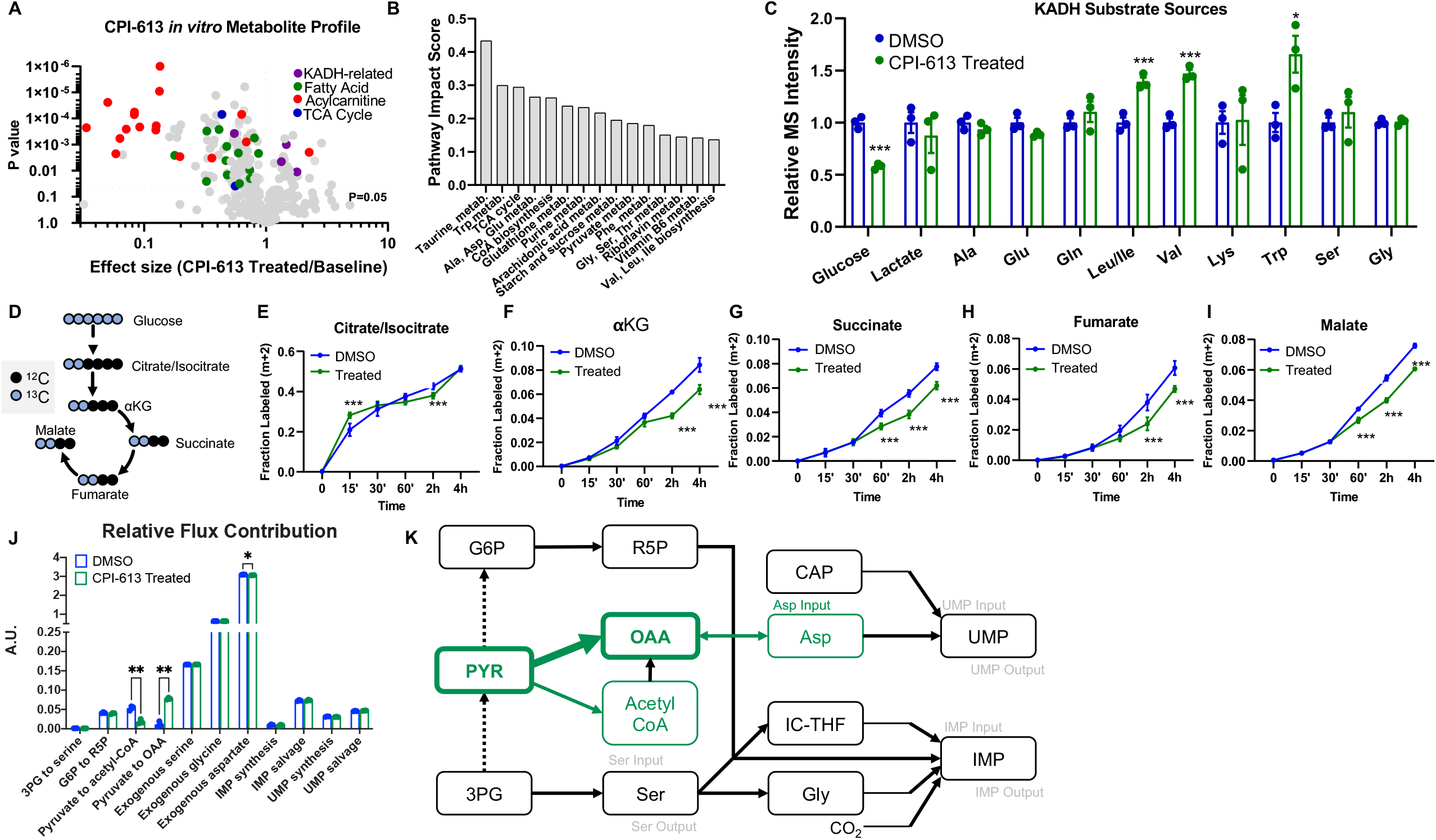
CPI-613 reduces KADH activity and mitochondrial metabolic flux in human cells. (a) Volcano plot of metabolites in response to cells treated with 130μM CPI-613 for 24h relative to DMSO control. Data are mean of three biological replicates. Purple dots, ketoacid dehydrogenase (KADH)-related metabolites; Green dots, fatty acids; Red dots, acylcarnitines; Blue dots, TCA cycle metabolites. These dots are metabolites that significantly changed between baseline and treated patients (p<0.05). (b) Network-based pathway analysis of all statistically significant metabolites altered by CPI-613 treatment of cells. (c) Relative mass spectrometry (MS) peak intensities of ketoacid dehydrogenase (KADH) substrate sources in cells treated with DMSO control or 130μM CPI-613 for 24h. Data are from three biological replicates, and error bars represent s.e.m. *P* < 0.05 [*], *P* < 0.005 [***], unpaired Student’s *t*-test. Ala, alanine; Trp, tryptophan; Lys, lysine; Glu, glutamate; Gln, glutamine; Val, valine; Ile, isoleucine; Leu, leucine; Ser, serine; Gly, glycine. (d) Schematic of [U-^13^C] glucose labeling of tri-carboxylic acid cycle intermediates. alpha-ketogluarate, αKG. (e-i) Fraction of the m+2 isotopomers of citrate/isocitrate, αKG, succinate, fumarate, and malate in the presence (green) or absence (blue) of 130μM CPI-613. Data are the mean of three biological replicates, and error bars represent s.e.m. *P* < 0.005 [***], unpaired Student’s *t*-test. In all experiments, cells were pre-treated with DMSO control or 130μM CPI-613 for 24h followed by introduction of [U-^13^C] glucose-containing medium including 130μM CPI-613 or DMSO control for the indicated times. (j) Metabolic flux analysis of the relative flux contribution to the indicated pathways in cells under control (DMSO) or CPI-613 treated conditions. Data are the mean of three biological replicates, and error bars represent s.e.m. *P* < 0.05 [*], *P* < 0.01 [**] unpaired Student’s *t*-test. (k) Schematic representation of the metabolic network and the effects of CPI-613 treatment assessed by metabolic flux analysis. PYR, pyruvate; G6P, glucose-6-phosphate; 3PG, 3-phosphoglycerate; R5P, ribose-5-phosphate; OAA, oxaloacetate; Ser, serine; Gly, glycine; Asp, aspartate; CAP, carbamoyl phosphate; UMP, Uridine monophosphate; IMP, inosine monophosphate; CO2, carbon dioxide.

## DISCUSSION

Identifying metabolic vulnerability in cancer has long been considered a potential therapeutic approach, and substantial basic science has been underway to define the rationale. Difficulties have been due to many reasons including a lack of clinical grade compound availability, an incomplete understanding of metabolic activities that define the window enabling tolerability to health and efficacy against malignancy, as well as predictive knowledge of the contexts in which such a therapeutic strategy could be effective. Complicating matters are the heterogeneity, plasticity, and redundancy that metabolic requirements exhibit. Thus defining measurable variables that are a part of mechanisms for when a particular metabolic state would be susceptible has been limited in the case of metabolic therapeutics.

The findings in this study show that specific measurable metabolic statuses related to the mechanistic target of action of CPI-613, ketoacid dehydrogenase enzyme complexes, can be used to determine patient response. We established in humans that a clinically active compound can engage its target with defined metabolic consequences related to its biochemical mechanism at the tumor site. Using metabolomics, we were able to show that metabolism consistent with targeting the mitochondria and the TCA cycle and related pathways could be observed in patients treated with the lipoic acid analog CPI-613. These effects were found systemically but became more pronounced at the tumor site, especially in responsive patients that exhibited gross alterations to TCA cycle activity and mitochondrial metabolism. This led to the development of a machine learning classifier based on three key pieces of mitochondrial biology – aspartate, acylcarnitine, and dihydroorotate metabolism. These metabolites are related to mitochondrial redox, fatty acid and nucleotide metabolism.

To our knowledge, this study represents the first in its kind to develop a predictive, mechanistic understanding of a metabolic therapeutic in human cancer that has shown high promise in early phase clinical trials. Additional mitochondrial metabolism inhibitors^8,24^ are currently in early phase clinical trials for treating cancer, as are other pre-clinical studies involving metabolic therapeutics^7,9,25^. Whether the concepts established in this study may be generalizable to other metabolic therapeutics in cancer is unknown but of interest for further study. Also, whether these results will extrapolate to the larger and randomized Phase III trials that are currently ongoing is unknown.

Finally, emerging work has now established that specific targeted alterations in diet can have a substantial impact on tumor metabolism and the efficacy of cancer therapies^26,27,28^. Thus, it will be interesting to explore how changes to the exogenous lipid or ketoacid dehydrogenase substrate source composition such as the availability of dietary branched-chain amino acids or a ketogenic diet may alter the response to CPI-613 treatment.

## METHODS

### Patient information

Blood and bone marrow plasma samples were obtained from Phase I and II Studies (NCT01768897, NCT02484391) of CPI-613 in combination with high dose cytarabine and mitoxantrone for relapsed or refractory Acute Myeloid Leukemia^2^ conducted at Wake Forest Baptist Health. All samples were obtained following signed informed consent and all protocols were approved by the Wake Forest Institutional Review Board. See Supplementary Table 1 for detailed patient information.

### Metabolite Extraction

Polar metabolite extraction from blood plasma and bone marrow plasma was performed as previously described^17^. Briefly, 20μL of blood plasma or bone marrow plasma was added to 80μL of ice-cold water, vortexed rigorously for one minute, and placed on ice for two minutes; then, 400μL of methanol (pre-chilled -80°C for 2 hours) was added per tube and vortexed rigorously again for one minute. Metabolite extractions were then centrifuged at 20,000xg at 4°C for 10 minutes, and the solvent in each sample was transferred to a new microcentrifuge tube before being evaporated using a speed vacuum. For polar metabolite analysis, the evaporated cell extracts were reconstituted by first dissolving in 15μL water and then diluted with 15μL methanol/acetonitrile (1:1 v/v). Samples were then centrifuged at 20,000xg at 4°C for 10 minutes, and the supernatants were transferred to LC vials prior to HPLC injection (3μL).

### High Performance Liquid Chromatography

An XBridge amide column (100 × 2.1mm i.d., 3.5μm; Waters) on a Dionex Ultimate 3000 UHPLC (at room temperature) was used for compound separation. The mobile phases were A) water with 5mM ammonium acetate, pH 6.9 and B) 100% acetonitrile with the following linear gradient: 0 min, 85% B; 1.5 min, 85% B; 5.5 min, 35% B; 10 min, 35% B; 10.5 min, 10% B; 12.5, 10% B; 13.5 min, 85% B; 20 min, 85% B. The flow rate was 0.15 ml/min from 0 to 5.5 min, 0.17 ml/min from 6.9 to 10.5 min, 0.3 ml/min from 10.6 to 17.9 min, and 0.15 ml/min from 18 to 20 min. Solvents (LC-MS grade) were purchased from Fischer Scientific.

### Mass Spectrometry

Mass spectrometry was performed using the Q Exactive plus (Thermo Scientific) instrument equipped with a heated electrospray ionization probe (HESI) as previously described^29^. The following parameters were used: evaporation temperature, 120°C; sheath gas, 30; auxiliary gas, 10; sweep gas, 3; spray voltage, 3.6 kV for positive mode and 2.5 kV for negative mode. The capillary temperature was set to 320°C, and S lens was 55. The resolution was set at 70,000. The maximum injection time was 200ms. The automated gain control was targeted at 3×10^6^ ions. The scan range used was 70 to 900 (m/z).

### Metabolite Peak Extraction and Data Analysis

Sieve 2.0 software (Thermo Scientific) was used to process raw peak data with peak alignment and detection performed according to the manufacturer’s protocol. The method “peak alignment and frame extraction” was applied for targeted metabolite analysis with an input file of theoretical m/z and detected retention times. The m/z width was set to 5 ppm. An output file was obtained after data processing that included detected m/z and relative intensity for each sample. To determine metabolic pathway impacts, MetaboAnalyst pathway analysis (www.metaboanalyst.ca) was used with the following parameters: Over Representation Analysis-Hypergeometric Test; Pathway Topology Analysis-Relative-betweenness Centrality. Partial least squares-discriminant analysis (PLS-DA) was used to determine patient variance between baseline and treated groups. For all box and whisker plots, the upper and lower whisker represents the maximum and minimum value, respectively, and the line represents the median value. Individual data points are juxtaposed on all box plots and bar graphs. All box plots, bar graphs, and volcano plots were generated using GraphPad Prism 8.

### Cell Culture

HCT116 cells were purchased from ATCC, authenticated by their source using STR-profiling, and tested negative for mycoplasma contamination. Cells were maintained in RPMI 1640 (GIBCO) supplemented with 10% heat-inactivated fetal bovine serum (Sigma, F2442) and 100 U/mL penicillin 100 mg/mL streptomycin (GIBCO). Cells were cultured in a 37°C, 5% CO_2_ atmosphere. Dialyzed fetal bovine serum (Thermo Fisher Scientific, 88440) was substituted for fetal bovine serum upon administration of the isotope-containing medium used in kinetic flux profiling experiments. U-^13^C-glucose was purchased from Cambridge Isotope Laboratories (CLM-1396-10).

### Kinetic flux profiling and metabolic flux analysis

Kinetic flux profiling and metabolic flux analysis was conducted as described previously^23^. For all experiments, cells were pre-treated with DMSO control or 130μM CPI-613 for 24h followed by introduction of [U-^13^C] glucose-containing medium including DMSO control or 130μM CPI-613 prior to metabolite extraction at the indicated times. Metabolic flux analysis was calculated from the 1 hour post-[U-^13^C] glucose-containing medium including administration time point.

### Machine Learning

For each metabolite, the mutual information coefficient (MIC)^30^ was calculated between the metabolite’s raw intensity values before treatment and the binary treatment response vector. Specifically, the MIC algorithm bins the data using a 2D grid of varying row and column sizes, computes the empirical distribution, and returns the maximum mutual information over a large number of grid configurations. The calculations were performed with the *mine* function of the *minerva* R package^31^. Metabolites with MIC values in the top 20 percent were selected. To predict response to treatment, a logistic regression model with the ridge constraint was fit from a given set of predictor metabolites using the *glmnet* R package^32^. The pre-treatment intensity values of the metabolites were first z-score normalized before being fed into the model. To predict the probability that a specific patient would respond to treatment, that patient’s data was held out, and a training set was constructed from the remaining patients in the dataset. A leave-one-out cross validation scheme was used to optimize for the ridge penalty of a logistic regression model using the training set. The model that minimized the misclassification error was then used to predict the treatment response probability for the hold-out patient sample. Repeating for every patient, the prediction scores were aggregated and compared to the ground truth labels. The true and false positive rates over the entire dataset were then computed to construct the ROC curves.

### Statistics

Patient sample size was indicated in the figure legends, and all cell culture experiments contained three biological replicates. Paired or unpaired Student’s *t*-tests were used to determine statistical significance of differences between means (*P* < 0.05 [*], *P* < 0.01 [**], *P* < 0.005 [***]) as indicated in the figure legends.

## Data Availability

All source code generated for data analysis and metabolomics data sets are available at https://github.com/LocasaleLab/Locasale-lab-2020-lipoic-acid-analog-study.

## ACKNOWLEDGEMENTS

We thank members of the Locasale laboratory for helpful discussions and comments on the manuscript. Support from the National Institutes of Health (R01CA193256 to J.W.L., R01 CA197991-05 to TSP) and the American Cancer Society (129832-RSG-16-214-01-TBE to J.W.L. and 131615-PF-17-210-01-TBE to M.A.R.) are gratefully acknowledged.

## AUTHOR CONTRIBUTIONS

M.A.R., T.S.P., and J.W.L. conceived the study and designed experiments. T.S.P. oversaw the Phase 1 and 2 CPI-613 (Devimistat) in combination with high dose cytarabine and mitoxantrone clinical trials and sample preservation. M.A.R. and S.B. performed the majority of the experiments. M.A.R. and J.W.L. wrote the manuscript. M.A.R., S.B., Z.X. and P.M. analyzed data. Z.D. and S.L. developed the metabolic flux analysis model and analyzed data. J.L. assisted with metabolomics methods and analyzed data.

## FIGURE LEGENDS

**Supplementary Figure 1.**
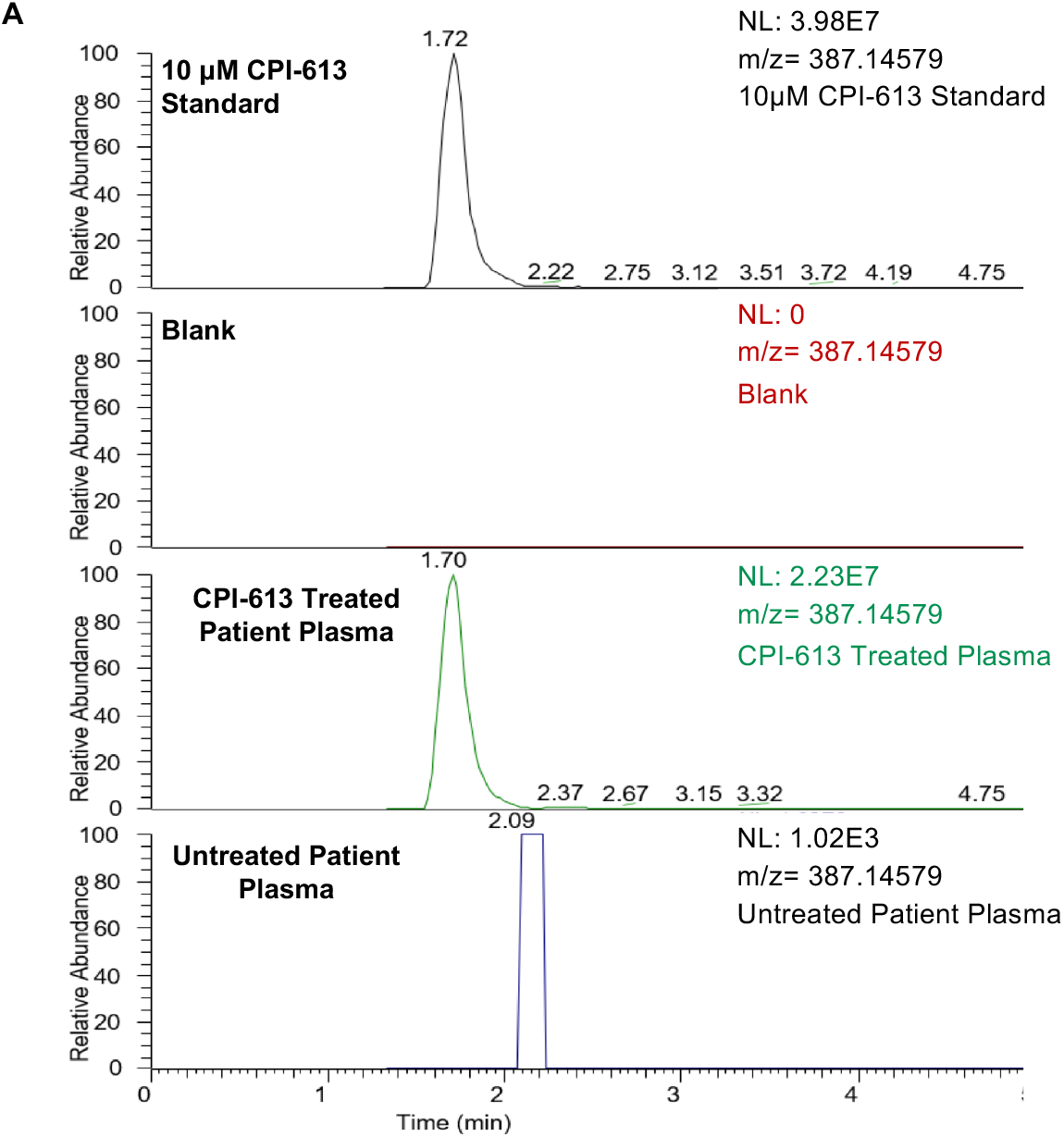
Chromatograms of CPI-613 MS peak in patient samples and standard. (a) Chromatograms of 10μM pure CPI-613 compound (top), solvent only blank (second from top) AML patient blood plasma after CPI-613 treatment (second from bottom) and AML patient blood plasma at baseline (bottom).

**Supplementary Figure 2.**
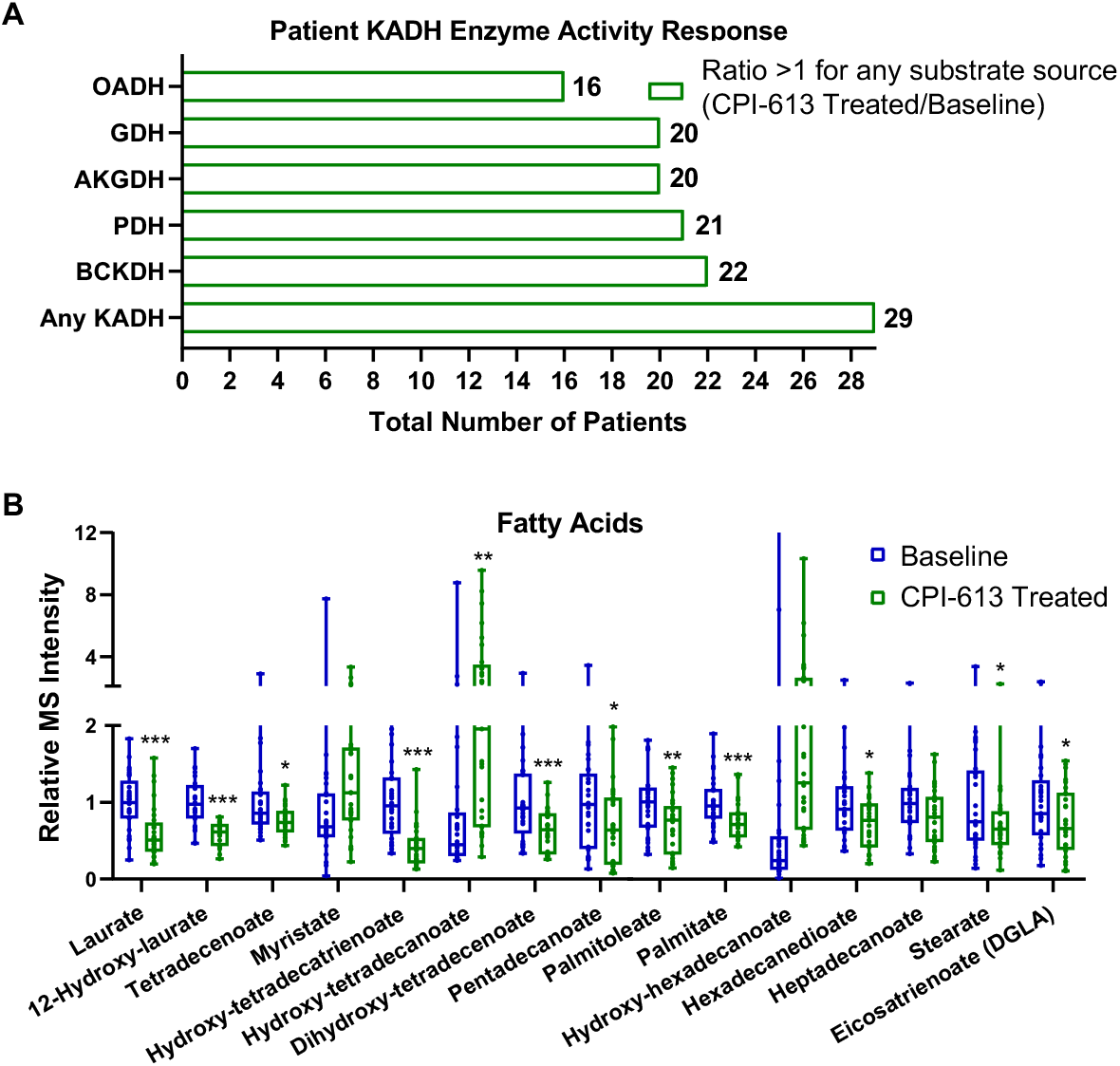
Blood plasma fatty acid profile in treated patients. (a) Total number of patients [out of 29] with reduced KADH enzyme activity, as indicated, after CPI-613 treatment as determined by substrate source accumulation calculated in Fig. 2c. (b) Relative mass spectrometry (MS) peak intensities of fatty acids in AML patient blood plasma at baseline and after CPI-613 treatment. The upper and lower whiskers of the box plots are the maximum and minimum values, respectively, and the bars are the median value. Data are from 29 AML patients, and individual values are plotted. *P* < 0.05 [*], *P* < 0.01 [**], *P* < 0.005 [***], paired Student’s *t*-test.

**Supplementary Figure 3.**
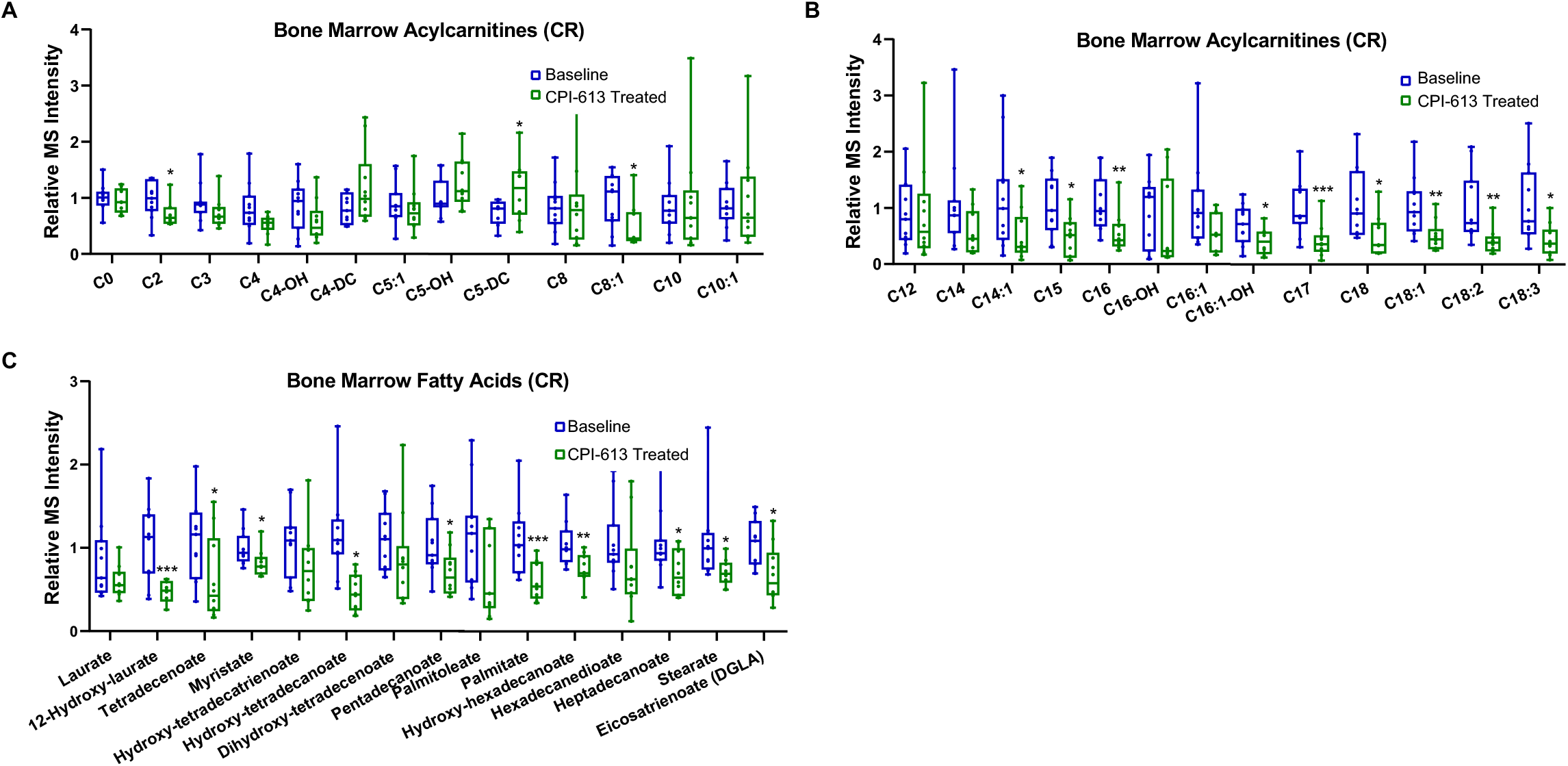
Bone marrow acylcarnitine and FA profiles in AML patients with CR. (a,b) Relative mass spectrometry (MS) peak intensities of acylcarnitines in complete remission (CR) AML patient bone marrow plasma at baseline and after CPI-613 treatment. The upper and lower whiskers of the box plots are the maximum and minimum values, respectively, and the bars are the median value. Data are from CR AML patients (Baseline n=11 patients; CPI-613 treated n=10 patients), and individual values are plotted. *P* < 0.05 [*], *P* < 0.01 [**], *P* < 0.005 [***], unpaired Student’s *t*-test. (c) Relative mass spectrometry (MS) peak intensities of fatty acids in complete remission (CR) AML patient bone marrow plasma at baseline and after CPI-613 treatment. The upper and lower whiskers of the box plots are the maximum and minimum values, respectively, and the bars are the median value. Data are from CR AML patients (Baseline n=11 patients; CPI-613 treated n=10 patients), and individual values are plotted. *P* < 0.05 [*], *P* < 0.01 [**], *P* < 0.005 [***], unpaired Student’s *t*-test.

**Supplementary Figure 4.**
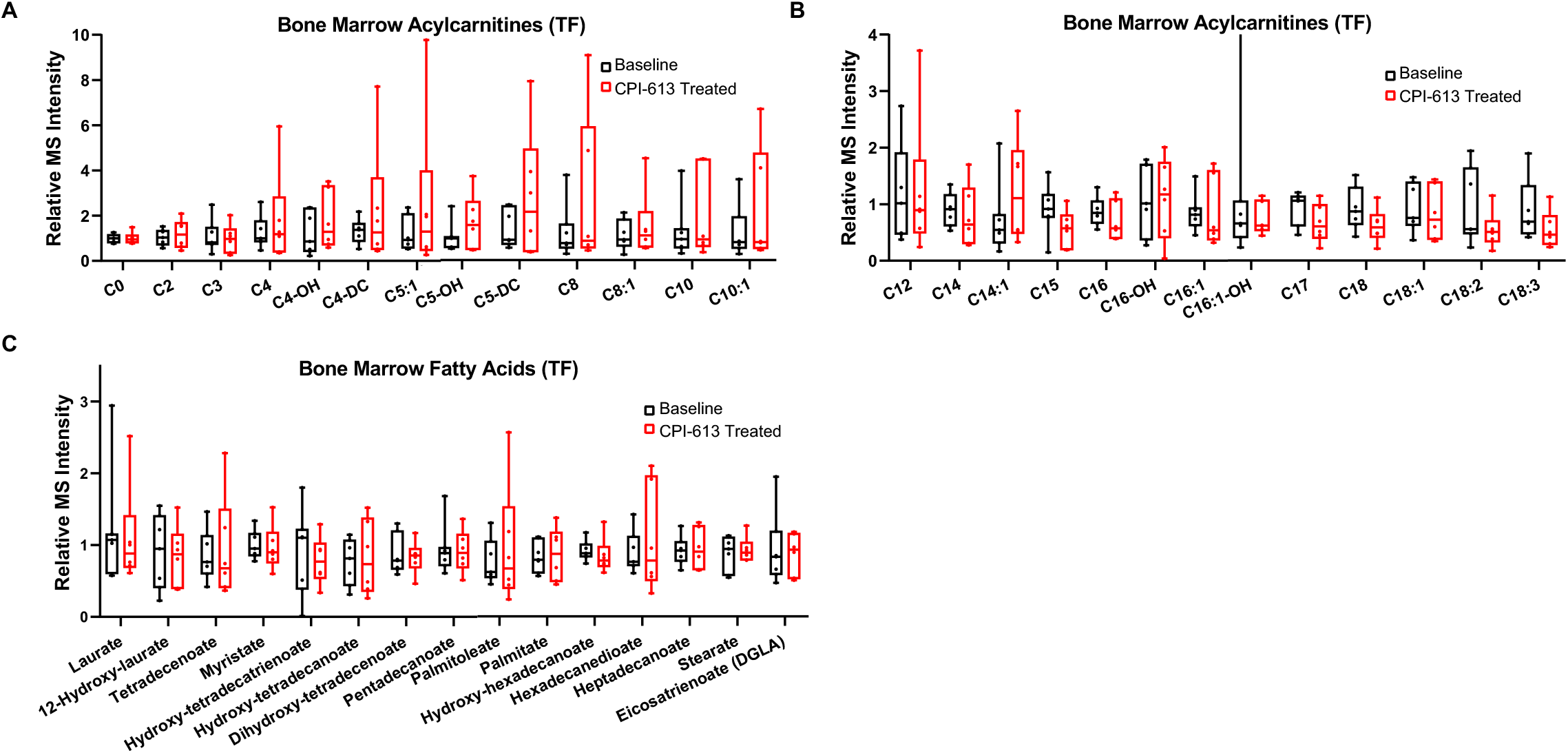
Bone marrow acylcarnitine and FA profiles in AML patients with TF. (a,b) Relative mass spectrometry (MS) peak intensities of acylcarnitines in treatment failure (TF) AML patient bone marrow plasma at baseline and after CPI-613 treatment. The upper and lower whiskers of the box plots are the maximum and minimum values, respectively, and the bars are the median value. Data are from AML patients (Baseline n=7 patients; CPI-613 treated n=6 patients). (c) Relative mass spectrometry (MS) peak intensities of fatty acids in treatment failure (TF) AML patient bone marrow plasma at baseline and after CPI-613 treatment. The upper and lower whiskers of the box plots are the maximum and minimum values, respectively, and the bars are the median value. Data are from AML patients (Baseline n=7 patients; CPI-613 treated n=6 patients).

**Supplementary Figure 5.**
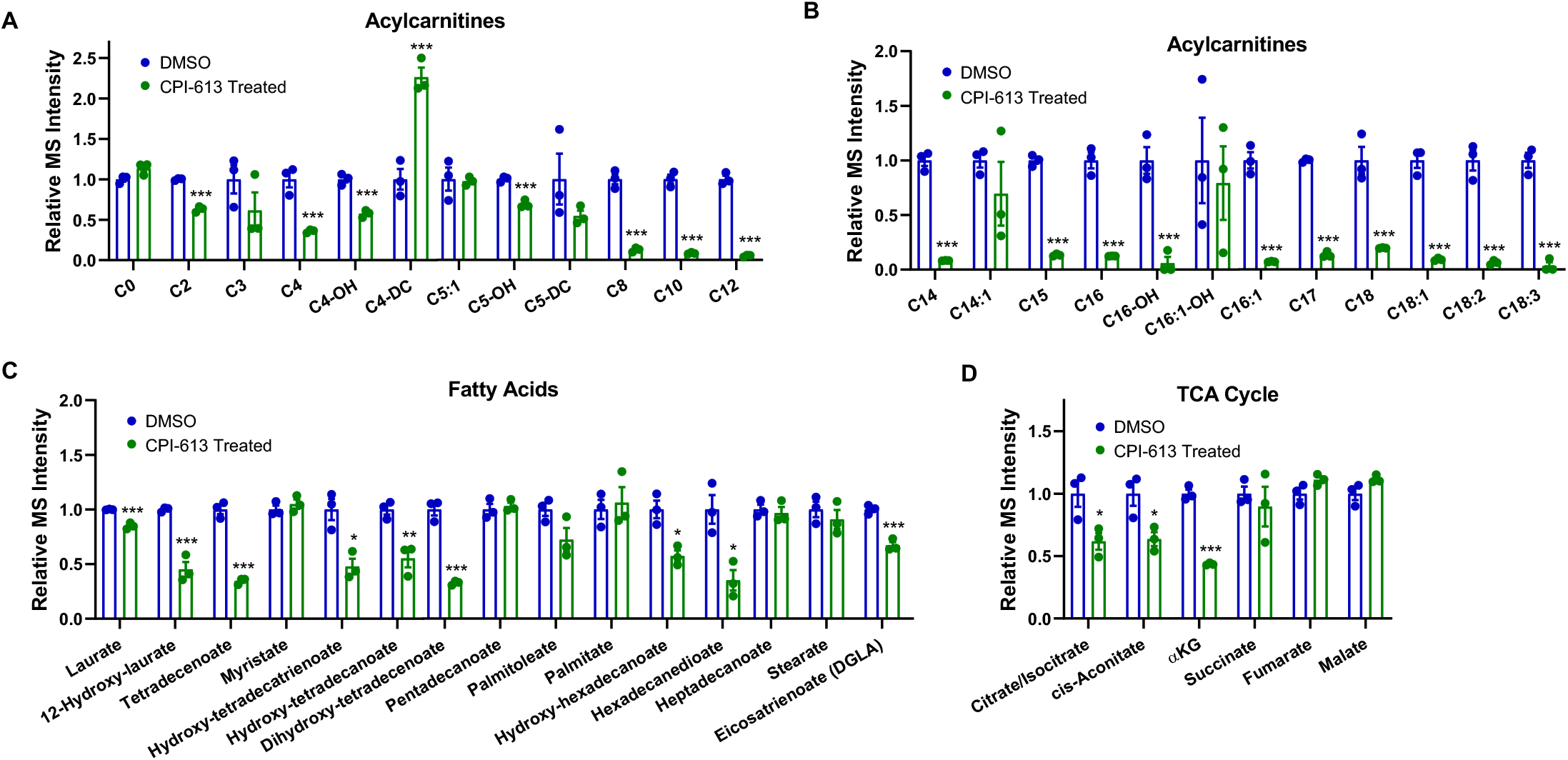
Cultured cells response to CPI-613 treatment. (a,b) Relative mass spectrometry (MS) peak intensities of acylcarnitines in cells treated with DMSO control or 130μM CPI-613 for 24h. Data are from three biological replicates, and error bars represent s.e.m. *P* < 0.005 [***], unpaired Student’s *t*-test. (c) Relative mass spectrometry (MS) peak intensities of fatty acids in cells treated with DMSO control or 130μM CPI-613 for 24h. Data are from three biological replicates, and error bars represent s.e.m. *P* < 0.05 [*], *P* < 0.01 [**], *P* < 0.005 [***], unpaired Student’s *t*-test. (d) Relative mass spectrometry (MS) peak intensities of tri-carboxylic acid (TCA) cycle metabolites in cells treated with DMSO control or 130μM CPI-613 for 24h. Data are from three biological replicates, and error bars represent s.e.m. *P* < 0.05 [*], *P* < 0.005 [***], unpaired Student’s *t*-test. alpha-ketoglutarate, αKG.

